# The Virtual Kitchen Challenge–Version 2: Validation of a Digital Assessment of Everyday Function in Older Adults

**DOI:** 10.1101/2025.08.12.25333444

**Authors:** Marina Kaplan, Moira McKniff, Stephanie M. Simone, Molly B. Tassoni, Katherine Hackett, Sophia Holmqvist, Rachel E. Mis, Kimberly Halberstadter, Riya Chaturvedi, Melissa Rosahl, Giuliana Vallecorsa, Mijail D. Serruya, Deborah A.G. Drabick, Takehiko Yamaguchi, Tania Giovannetti

## Abstract

**Background:** Conventional methods of functional assessment include subjective self/informant-report, which may be biased by personal characteristics, cognitive abilities, and lack of standardization (e.g., influenced by idiosyncratic task demands). Traditional performance-based assessments offer some advantages over self/informant reports, but they are time consuming to administer and score.

**Objective:** To evaluate the validity and reliability of the Virtual Kitchen Challenge -Version 2 (VKC-2), an objective, standardized, and highly efficient alternative to current functional assessments for older adults across the spectrum of cognitive aging, from preclinical to mild dementia.

**Methods:** 236 community-dwelling diverse older adults completed a comprehensive neuropsychological evaluation to classify their cognitive abilities as healthy, mild cognitive impairment or mild dementia, after adjustment of demographic variables (age, education, sex, estimated IQ). Participants were administered the VKC-2, which required completion of two everyday tasks (breakfast, lunch) in a virtual kitchen using a touch-screen interface to select objects and sequence steps. Automated scoring reflected completion time and performance efficiency (e.g., number of screen interactions, % time spent off screen, interactions with distractor objects). Participants also completed the VKC-2 tasks using real objects (Real Kitchen) and questionnaires of everyday function. Informants for 219 participants completed questionnaires regarding everyday function. A subsample of participants (n = 143) performed the VKC-2 again in a second session 4-6 weeks after the baseline for retest analyses. Analyses evaluated construct and convergent validity and retest and internal reliability of VKC-2 automated scores.

**Results:** Construct validity was supported by ANCOVA results showing participants with healthy cognition obtained significantly better VKC-2 scores than participants with cognitive impairment (all ps < .001), even after controlling for demographics and general computer visuomotor dexterity. Convergent validity was supported by significant correlations between VKC-2 scores and performance on the Real Kitchen (r values = -.58 to .64, ps < .001), conventional cognitive test scores (r values = -.50 to -.22, ps < .001), and to self and informant questionnaires evaluating everyday function (r values = .25 to .43, ps < .001). Retest reliability was strong as evidenced by high intraclass correlations between VKC-2 scores across the two timepoints (κ = 0.27, p < .001). Reliability improved in analyses including only participants who reported no change in cognitive status between time 1 and time 2 (n=123). Spearman-Brown correlations showed acceptable to good internal consistency between the VKC-2 tasks (breakfast, lunch) for all scores (.77 to .81) supporting the use of total scores.

**Conclusions:** The VKC-2 is an efficient, valid, and sensitive measure of everyday function for diverse older adults that may be used for large scale longitudinal studies, clinical trials, and clinical assessments. The VKC-2 holds promise to improve the status quo of functional assessment in aging particularly when informants are unavailable or unreliable.

---

As the U.S. population ages and interventions for Alzheimer’s disease (AD) and AD related dementias (ADRD) become available[1], highly sensitive, objective and efficient measures of functional abilities are needed for multiple purposes. Mild functional difficulties are one of the strongest predictors of future cognitive decline and dementia[2–5]; thus, accurate measurement of functional ability will improve prediction of prognosis and identify the need for early intervention. Given that functional ability level is often the criterion that distinguishes mild cognitive impairment (MCI) from mild dementia, accurate assessment is critical for diagnostic decision-making[6,7]. According to the Food and Drug Administration (FDA), the approval of pharmacological treatments for dementia, even at the very early, pre-symptomatic stage, is contingent on demonstrating gains on meaningful measures of functioning[8]. Recently approved treatments have relied on composite measures such as the Clinical Dementia Rating–Sum of Boxes and the integrated Alzheimer’s Disease Rating Scale, but these measures require specialized training, are not readily deployable in typical clinical settings, and lack sensitivity to the earliest functional changes[9,10]. There exists a critical need for sensitive and efficient functional assessment tools that are clinically meaningful, psychometrically sound, and practically implementable across diverse healthcare settings [11,12]. We developed a non-immersive virtual reality (VR) measure, the Virtual Kitchen Challenge -Version 2 (VKC-2), an objective, sensitive, efficient, and theoretically based tool for assessment of everyday function in older adults to address the gaps in current functional assessments. Here we report results on the VKC-2 validity and reliability in racially diverse, community-dwelling older adults with healthy cognition, MCI, or mild dementia.

In practice, self/informant reports of everyday function, which are the current standard method for functional assessment, are easy to administer and score, and when used with reliable, observant, and knowledgeable reporters, they generate very useful information about how a person is functioning in everyday life[13–15]. In many circumstances, however, the accuracy of self and informant report is uncertain. Their subjective nature makes them prone to over- or under-reporting due to faulty cognitive abilities, psychological factors (e.g., denial, depression, burden), or cultural beliefs[16]. Informant report is often unavailable, such as for individuals without a living spouse, nearby family members, or close friends. Even when available and willing, informants may have limited opportunities for observation of daily functioning and may lack knowledge, particularly when functional difficulties are mild and may be masked by compensatory behaviors[17,18].

Another limitation of questionnaires is that older adults vary widely in the activities that they perform and the contexts in which they perform them. For example, informant reported difficulties with medication management may be profoundly different for an older adult managing a single prescription and maintains a small, organized home with her spouse versus an older adult who taking dozens of medications living in a large, cluttered house[19]. However, given identical clinical presentations and cognitive test scores suggesting mild cognitive decline, the latter patient would likely to be diagnosed with clinical dementia if she were unable to independently manage her medications. Failure to account for task complexity and contexts confounds the informant report of everyday function and precludes clear comparisons of functional abilities across individuals.

Further, many questionnaires do not distinguish difficulties due to physical versus cognitive limitations[14], and if they do, it might be difficult for an informant to fully understand the nature of functional difficulties, particularly because physical and cognitive difficulties often co-occur[20–22]. Informant and self-reports also do not offer detailed characterization of types of functional difficulties due to varying underlying cognitive problems (e.g., slowing, disorganized actions vs. omission of crucial task steps), which could offer insight into interventions for improving function and reducing risk of future functional disability[23,24].

Performance-based measures of function address many of the limitations of questionnaires; they are objective, they standardize task complexity and context, and they allow for detailed analysis of behavior and systematic comparison across individuals. The Naturalistic Action Test (NAT), for example, is a performance-based test of everyday function with strong psychometric properties, normative data, and suggested cut scores for healthy cognition vs. MCI vs. mild dementia[23,25–35]. Scoring NAT performance for subtle inefficient errors, called micro-errors, has increased the sensitivity of NAT tasks for detecting mild difficulties with everyday tasks[35–38]. Results from performance-based tests, like the NAT with added sensitive scoring procedures, have demonstrated that 1) healthy older adults make more errors and require more time to complete everyday tasks than younger adults[36,37,39–43]; 2) people with MCI make more errors than healthy controls but fewer errors than individuals with dementia[32,35,44–46]; 3) the ability to accurately and efficiently perform everyday tasks is moderately correlated with performance on cognitive tests[27,28,35,37,47] and informant report of everyday function[27–29,31,35,48]. Together, these findings and others[37,49–52] suggest that standardized performance-based assessment of function is valid and reliable.

Despite their objectivity, validity, and potential for rich characterizations of function, current performance-based tests require extraordinary effort, limiting their implementation and scalability. Scoring, particularly scoring for subtle errors and inefficiencies, is time-intensive and requires video recording, detailed scoring instructions, and trained coders. Although some performance-based tests may be scored quickly as pass/fail without video recording[40,53], such gross measures are less sensitive to mild difficulties (i.e., MCI)[54], do not advance our understanding of the nature of functional problems[4,55,56], and/or still require considerable effort to administer. To streamline administration and scoring, a non-immersive, VR task called the Virtual Kitchen, modeled after the NAT, was developed. The original version of the Virtual Kitchen[57] required a mouse to move objects on a computer screen to complete a coffee making task. Results showed that people with dementia (n =24) accomplished fewer steps and made more errors than healthy controls (n = 32) on the Virtual Kitchen. Validity also was supported by significant correlations between the Virtual Kitchen scores and performance of real tasks, cognitive tests, and informant reports of functioning[57].

Our team developed a revision of the original Virtual Kitchen[57], called Virtual Kitchen Challenge (VKC), by implementing the following updates: 1) expanding the coffee task to include a more extensive breakfast; 2) adding a lunch task; 3) updating the graphics; and 4) transitioning from a mouse to a computer touchscreen to make interactions more natural[39]. We also added a brief training task to familiarize participants with the touchscreen interface. Automated scores were expanded to include measures computed based on interactions with the touchscreen to increase sensitivity (i.e., number of screen interactions, etc.). Preliminary results from a sample of 14 older adults and 21 younger adults demonstrated validity and good internal consistency[39]. Subsequent studies showed the validity of the VKC automated scores against conventional cognitive tests in young adults [52]and against neuroimaging markers of cerebral vascular disease (white matter hyperintensities) in a small sample of community-dwelling older adults[48].

In this paper we present the psychometric properties of the automated scores from the most recent update of the Virtual Kitchen, the Virtual Kitchen Challenge-Version 2 (VKC-2). This version includes enhanced graphics and a more extensive basic familiarization task for practice and to obtain a score of participants’ basic digital visuomotor dexterity that may be used as a control measure. We evaluated construct and convergent validity as well as retest and internal reliability of the VKC-2 in a large community-based sample of racially diverse older adults with healthy cognition, MCI, or mild dementia. Construct validity of the VKC-2 was evaluated in a known group comparison of automated scores (healthy cognition vs. MCI vs. mild dementia). Convergent validity was evaluated with correlations between automated VKC measures and performance on the real versions of the VKC-2 tasks (Real Kitchen), demographically adjusted cognitive test scores, and conventional self/informant questionnaires of everyday function. Retest reliability was evaluated over a period of 4-6 weeks. Internal consistency was evaluated for the two VKC-2 tasks (breakfast, lunch).

## Methods

### Participants

Participants were recruited for an observational, longitudinal psychometric study designed to evaluate the psychometric properties of the VKC-2 (n = 217; R01AG062503) or for a separate, smaller study on activity tracking (n = 20; F31AG089944). Procedures for the baseline visit of both studies were the same, designed and conducted in accordance with the Helsinki Declaration, and approved by the Institutional Review Board at Temple University (IRB protocol 23116 & 29712). All participants signed informed consent forms, were compensated for their participation ($50 USD for participants per session; $25 USD for informant per session), and were assigned study numbers to protect their privacy when storing research records. At the end of the study, participants also were offered a research report with their cognitive tests scores, if interested.

### Participants

All participants were recruited from community outreach events, fliers, and referrals from neurology departments in Philadelphia, Pennsylvania from September 2020 through June of 2025. Inclusion/exclusion criteria were screened by phone, with only minor differences between the two studies. For both studies participants were excluded for the following reasons: lifetime history of severe psychiatric disorder (e.g., schizophrenia, bipolar disorder), nervous system infections, or disorders (e.g., epilepsy, brain tumor); current metabolic or systemic disorders (e.g., B12 deficiency, renal failure, cancer); current moderate-severe depression; current moderate-severe anxiety symptoms; severe sensory deficits that would preclude visual detection or identification of common everyday objects used in the study or the inability to hear the task directions (e.g., blindness, total hearing loss); severe motor weakness that would preclude the use of everyday objects (e.g., severe deformities or paralysis of both upper extremities); intellectual disability; not being a fluent English speaker. The inclusion/exclusion criteria for the larger study required participants to be at least 65 years old and have an available informant who could serve as a study partner. Informants were screened by phone for the following eligibility criteria: 18 years of age or older; fluent English speaker; available and willing to complete study questionnaires in person, by phone, or online; has daily contact with the participant and reports knowledge of the participant’s daily functioning. Inclusion/exclusion for the second, smaller study required participants to be at least 55 years old and did not require a study informant/partner.

### Procedures

At the baseline visit (session 1), participants (N = 237) completed informed consent, cognitive testing, the VKC-2, the real version of the VKC-2 (i.e., Real Kitchen), and questionnaires regarding demographic information and their familiarity with the tasks used in the VKC-2. Order of the Real Kitchen and VKC-2 was counterbalanced across participants to control for order effects. At session 1 informants completed questionnaires in-person, online, or at home by mail. After reaching our target sample size (n ≈140) for retest reliability analyses (June 2024), participants were no longer requested to return for a second session 4-6 weeks after session 1 [58]. A total of 143 participants completed session 2, which included a brief interview (for both the participant and informant) regarding changes in cognition or health status (e.g., medication changes, falls, illnesses, hospitalizations, etc.) since session 1 as well as repeat administration of the VKC-2 and Real Kitchen.

### Measures

#### Cognitive Tests

Cognitive tests were administered to characterize the sample, classify participants according to their cognitive status, and evaluate the convergent validity of the VKC-2. The cognitive testing protocol is described in Table 1. The protocol included two tests from four different cognitive domains to characterize participants according to Jak/Bondi actuarial criteria[59,60] and clinical criteria originally proposed by Petersen[6] and McKhann[61]. Specific tests had normative data from the Calibrated Neuropsychological Normative System(CNNS)[62] to enable raw score adjustments for sex, age, education, and estimated IQ. Such demographic adjustments are critical for confirming group membership in a diverse sample of older adults. Further details on how tests were used for classifying cognitive abilities are provided in Supplemental Materials. For analysis of VKC-2 convergent validity, composite scores were computed by averaging demographically adjusted T scores from tests within each domain (Episodic Memory, Language, etc.). A global cognitive composite was modeled after the Knight-Preclinical Alzheimer Cognitive Composite (modified [m]Knight-PACC)[63], which has been shown to be sensitive for detecting early cognitive change due to neurodegenerative disease.

**Table 1.** Cognitive tests administered at session 1.

| Cognitive domain | Test | Score(s) | Reference |
| --- | --- | --- | --- |
| Premorbid intellectual functioning (IQ) | Hopkins Reading Test | Estimated IQ | Schretlen, 2009[64] |
| Global cognitive status | Mini-Mental State Examination | Total correct | Folstein et al., 1975[65] |
| Episodic memory | *†Hopkins Verbal Learning Test–Revised | Delayed free recall, total correct | Brandt & Benedict, 2001[66] |
|  | *Brief Visual Memory Test–Revised | Delayed free recall, total correct | Benedict et al., 1996[67] |
| Language | *†Category (Animal) Fluency | Total correct | Schretlen, 2010[62] |
|  | *Boston Naming Test–30 item | Total correct | Goodglass & Kaplan, 1983[68] |
| Executive function | *†Trail Making Test–Part B | Completion time | Reitan, 1958[69] |
|  | *Digit Span Backward | Longest span | Wechsler, 1987[70] |
| Processing speed | *†Salthouse Letter Comparison | Total correct | Salthouse, 1991[71] |
|  | *†Salthouse Pattern Comparison | Total correct | Salthouse, 1991[71] |
| Attention | *Digit Span Forward | Longest span | Wechsler, 1987[70] |
|  | *Trail Making Test A | Completion time | Reitan, 1958[69] |

*Note.* \**T*-scores from these tests were used for Healthy vs. MCI vs. dementia classification. †*T*-scores from these tests were averaged to compute the mKnight-PACC.

#### Virtual Kitchen Challenge-Version 2 (VKC-2)

The VKC-2 is a non-immersive virtual reality test of everyday function that requires participants to complete two everyday tasks (breakfast, lunch) by moving virtual objects using a touch-screen[39,57]. The VKC-2 tasks and objects were modeled after the Naturalistic Action Test (NAT)[25], an extensively studied and theoretically based performance-based test of everyday function that involves completion of familiar everyday tasks using real objects. The VKC-2 breakfast and lunch tasks were designed to be of comparable complexity and difficulty, with each task including thirteen target objects and four distractor objects.

For this study, the VKC-2 was administered on a MSI Creator Z16-A12UET laptop (12th Gen Intel® Core™ i9 Processor) with a 16” QHD+ (2560×1600), 120Hz, IPS-Level Touchscreen Display to maximize visibility and portability. Participants were instructed to use the index finger of their dominant hand to move and manipulate objects on the touchscreen.

The VKC-2 included three phases: Movement Familiarization, Task Training, and Test. See Figure 1 and below for more details.

**Figure 1.**
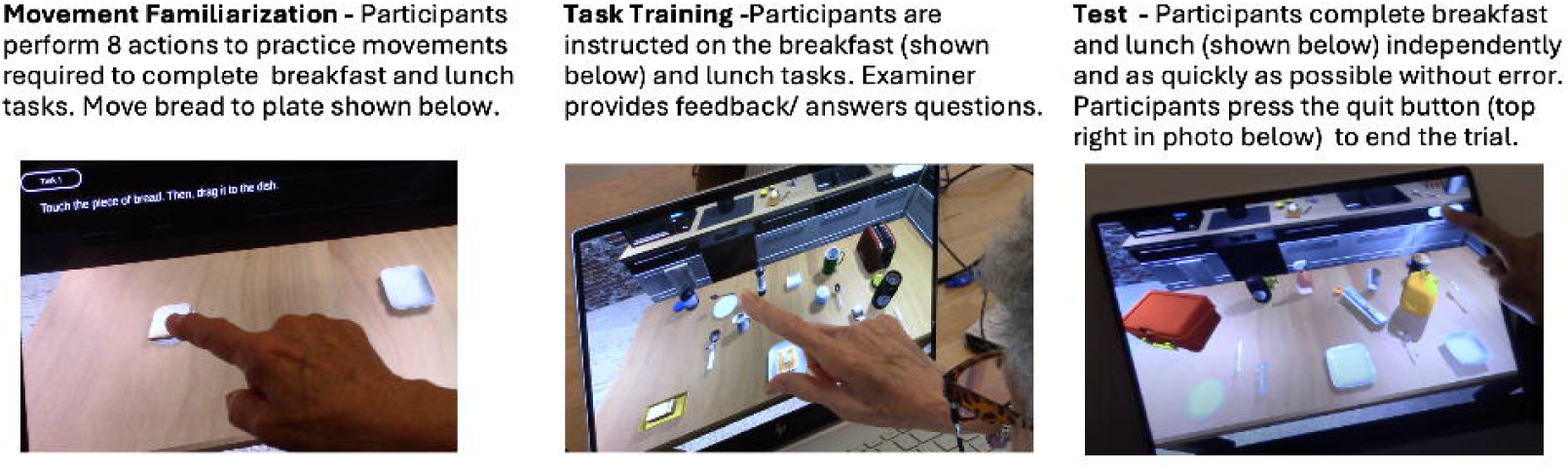
(Photos of participants completing each phase of the VKC-2)

##### VKC-2 Movement Familiarization

Participants were directed to perform eight simple movements to familiarize them with the basic touchscreen actions (e.g., tap, drag). The movement familiarization items included: 1) move bread to dish, 2) stir mug with spoon, 3) pour juice, 4) place thermos in lunch box, 5) spread jelly on bread, 6) wrap cookies in foil, 7) place bread in toaster, and 8) add sugar to mug. Participants first performed all the basic touchscreen actions with guidance of the examiner and the opportunity to ask questions and repeat each action as needed. Next, participants were asked to complete all eight trials independently as quickly and efficiently as possible. Completion time of the second, independent trial was computed as a measure of basic digital visuomotor dexterity (Digital Dexterity Score).

#### VKC-2 Task Training

The examiner reviewed written instructions that were presented on the computer screen for each task. Then, participants were asked to point to each of the target objects needed for each task. For example, training for the breakfast task included a direction to “point to all of the objects you will need for the toast” as the examiner named the object out loud (e.g., “bread,” “toaster,” etc.). Participants also were asked to point to each of the distractor objects and were told that they would not need to touch or use those objects. Participants then proceeded to the practice trials where they made the breakfast and lunch with prompting, cues, and error correction from the examiner. The examiner also answered questions to ensure that participants fully understood each task.

#### VKC-2 Test

Breakfast and lunch tasks were completed independently without feedback. Instructions regarding the task objectives that were reviewed during the practice trial were repeated (e.g., “pack a lunch for someone who wants a sandwich, snack and a drink”). Participants also were told to complete test trials as quickly as possible, without making errors, and using clear and precise movements. They were told to touch the quit button at the top right of the screen to end the trial (see Figure 1). Participants were asked to verbally repeat the directions before each task to ensure comprehension; instructions were repeated as often as needed before the participant initiated the task.

#### VKC-2 Test Automated Scores

Performance on the VKC-2 Test tasks (breakfast + lunch) were scored using data from the touchscreen and as described and validated in our pilot work with the original version of the VKC[39,48,52]:

1) Completion time (time) was recorded in seconds from the point at which the virtual kitchen screen appears (after instructions) until the time the quit button is pressed by the participant. Results from prior studies of the original VKC indicate that completion time differed significantly between older and younger participants and correlated with completion time on the Real Kitchen, cognitive tests of executive function and episodic memory[39], and neuroimaging markers of cerebrovascular disease[48].
2) Number of screen interactions (touches) included the number discrete instances the participant made contact with the computer touch screen. Touches were collected as a measure of performance efficiency, with fewer screen interactions reflecting more precise and deliberate actions. Results with the original VKC showed that older adults made significantly more touches than younger adults, with additional touches by older adults including both inefficient correct actions and errors. A higher number of touches was significantly associated with more total errors scored by trained coders who watched video recordings of the VKC performance. Additionally screen interactions were significantly associated with performance on the Real Kitchen and cognitive tests of executive function and episodic memory[39].
3) Percent of time off-screen (%off-screen) was the time spent working on the VKC-2 when the participant was not touching the screen; it was computed by subtracting the time spent touching the screen from the completion time and then dividing by the completion time and multiplying by 100. The percent of time off-screen also reflects performance efficiency. Pilot data from the original VKC[39] indicated that older adults spent a significantly higher percentage of their total time off-screen than younger adults. Correlations between %off-screen and human codes of the VKC performance suggested that higher %off-screen times were due to multiple factors, including slower planning, difficulties locating target objects, difficulty resolving competition for object selection, and misreaching toward the computer screen (i.e., micro-errors). Higher %off-screen times were significantly associated with more errors on the Real Kitchen and poorer scores on tests of executive function[52] and episodic memory[39] and neuroimaging markers of cerebrovascular disease[48].
4) Number of distractor object interactions (distractor interactions) include instances when a distractor object is touched and/or moved. Our pilot work in a sample of healthy older and younger adults indicated that distractor interactions occurred too infrequently for analyses[39], but they have not been studied in participants with cognitive impairment.

#### Real Kitchen

The Real Kitchen required participants to complete the breakfast and lunch tasks using real objects that were placed on a table See Figure 2. Instructions for the Real Kitchen were identical to the VKC-2, including the instruction to “press the quit button when finished.” In the Real Kitchen, the Quit Button was a piece of paper on the right side of the table that said “QUIT.” Real task objects were similar in appearance (color, shape) to the simulated objects in the VKC-2. Participants repeated the directions before each test trial to indicate comprehension; instructions were repeated as often as needed. Participants were video recorded; recordings were labeled using a code so human coders were unaware of participant classification and study session.

**Figure 2.**
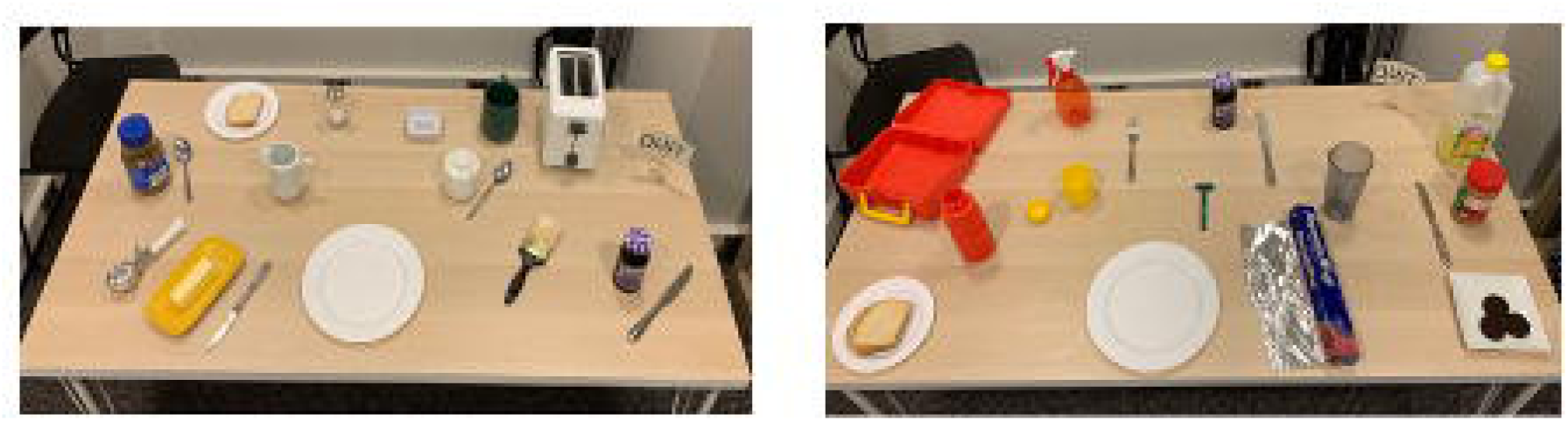
(Real Kitchen breakfast and lunch tasks)

Real Kitchen performance was scored according to detailed instructions using well validated scores and procedures. Real Kitchen Scores from a subset of the current sample have been published and show strong inter-rater reliability, significant differences between participants with healthy cognition versus cognitive impairment, and correlation with cognitive tests and self/informant-report of everyday function[35]. For the current study, the following Real Kitchen scores were used for validation (convergent validity) of the VKC-2 automated measures:

1) *Real Kitchen completion time* was recorded in seconds; and has been reliably coded by starting to time when the first step was initiated and ending when the participant touches the quit button. Prior work shows participants with greater cognitive impairment demonstrate longer completion times than participants with healthy cognition[35].
2) *Accomplishment* was coded for each completed step and is scored from 0-13 for the breakfast task and 0-20 for the lunch task. A total accomplishment score was computed (0-33), with higher scores reflecting more task steps accomplished.
3) *Total errors* were coded according to a taxonomy previously published in numerous studies with a range of clinical populations[25,72], showing validity and strong inter-rater reliability in people with stroke[73,74], dementia[27,28,30,47], MCI[26,32,33], and healthy controls [37,38,49], as well as a subset of participants from this sample[35]. The scoring taxonomy (see Supplemental Materials) includes overt errors (e.g., performing task steps in wrong sequence) and micro-errors (e.g., reaching toward a distractor object). In studies of dementia participants, total overt errors correlate with cognitive tests and informant reports of function. The micro-error category was added to improve detection of subtle, inefficient behaviors in healthy and MCI participants [35,37,38,49]. Because overt errors occur with relatively low frequency, they were combined with micro-errors to compute a total error score[35].
4) *Motor errors* were tracked separately from total errors. Motor errors involved instances where the correction action was performed with motor or spatial imprecision (e.g., spill coffee grinds, drop knife, etc.).

#### Participant Questionnaires

Participants completed a demographic form assessing age, sex, race, ethnicity, income, and education level and the following questionnaires:

1) Past Experience Scale[45,75] assessed familiarity with the breakfast and lunch tasks that comprise the VKC/Real Kitchen with 4 items (toast, coffee, sandwich, thermos) each rated on a scale from 0 (not at all familiar) to 4 (very familiar). Total Familiarity score ranged from 0 - 16 with higher scores reflecting greater familiarity. Frequency with which participants competed each task in their day-to-day life on average over the past 5-10 years also was rated for each item using a scale from 0 (never) to 4 (just about every day), with total scores ranging from 0 (never performed any of the tasks) to 16 (performed each task just about every day).
2) Functional Activity Questionnaire (FAQ[14]) instructions were modified to reflect only difficulties due to cognitive problems (not physical problems, fatigue, etc.) for 10 activities (e.g., preparing a balanced meal). Each activity is rated on a scale from 0 (performs normally) to 3 (dependent). Total FAQ scores range from 0 to 30 with higher scores reflecting greater dependence on others in everyday tasks due to cognitive difficulties.
3) Everyday Cognition Scale (ECog-12[13,76]) measures decline over the past 10 years in 12 everyday cognitive abilities (e.g., remembering where you have placed objects) on a scale from 1 (better or no change) to 4 (much worse all the time). Total scores reflect an average across all completed items and range from 1-4, with higher scores indicating a greater decline in everyday cognition.
4) Instrumental Activities of Daily Living - Compensation (IADL-C[15]) measures the need for assistance and compensatory strategies when performing 27 daily activities (e.g., prepare own meals). Each activity is rated on a scale from 1 (independent, no aid) to 8 (not able to complete activity anymore). The total score is the sum of all item responses, with a possible range from 27 (completely independent, no aid needed for any tasks) to 216 (no longer able to perform any task).

#### Informant Questionnaires

Informants completed questionnaires regarding their demographic information (age, education, etc.), their relationship with the participant (e.g., cohabitation, years known, hours in contact with the participant), and the participants’ everyday function, including the ECog[13,76], FAQ[14], and IADL-C[15]. Instructions and scoring for each of the questionnaires was the same as participant versions described above.

### Analysis Plan

Analyses were conducted using SPSS Version 29.0[77]. VKC-2 automated scores were examined for outliers and Winsorized at the 99^th^ and 1^st^ percentile. The VKC-2 distractor interaction score was dichotomized because few participants interacted with distractor objects (0

= no interactions with distractor objects; 1 = at least one interaction with a distractor object during completion of the VKC-2).

#### Construct Validity

VKC-2 automated scores were compared across groups that are known to differ in their functional ability level: healthy cognition, MCI, and mild dementia. Because the size of the dementia subgroup was relatively small (n =16), statistical analyses focused on differences between participants with healthy cognition vs. MCI. Participants with dementia were included for descriptive comparisons. One-way analyses of covariance (ANCOVA) were used to test group differences for each VKC-2 automated score after controlling for demographics: digital dexterity, time, touches, %off-screen. Group differences also were evaluated in ANCOVA models that also controlled for the digital dexterity score to determine whether significant group differences were explained by differences in basic visuomotor or computer abilities. Group differences on the dichotomized VKC-2 distractor interaction score was evaluated using Chi Square tests. Significant between group differences with moderate effect sizes (i.e., eta squared > .01; Phi (φ) coefficient > .30) were interpreted as support the construct validity of the VKC-2 automated scores.

Receiver Operating Characteristic (ROC) analyses comparing participant groups (healthy cognition vs. impaired cognition [MCI +dementia]; healthy cognition vs. MCI) were performed to identify cutoff values for each of the VKC-2 automated scores. Youden indices were used to identify cut scores that optimized sensitivity and specificity[78].

#### Convergent Validity

Correlations between the VKC-2 automated measures and the ability to perform tasks with real objects (Real Kitchen), demographically adjusted cognitive test scores of overall cognition and specific cognitive abilities and self/informant-reports of everyday functioning were performed to evaluate convergent validity. Pearson correlation coefficients were computed with the full sample. Spearman-rank order correlations were also performed and are included in Supplemental Materials. Significant and moderate level relations were interpreted as supporting the convergent validity of the VKC-2 automated scores.

#### Reliability

Retest reliability was assessed using Intraclass Correlation Coefficients (ICCs), calculated with a two-way mixed-effects model based on absolute agreement and average measures[79]. ICC values range from 0 to 1, with values above .75 generally indicating good reliability and values above .90 considered excellent[80]. Confidence intervals (95%) were computed for each ICC, and significance was determined using F-tests. Test–retest reliability for the distractor interaction score (dichotomous variable) was examined using Cohen’s kappa. Retest reliability was evaluated for the full sample who completed session 2 (n = 143) and for a subsample that reported no change in cognitive abilities since session 1 (n = 123/143). Internal consistency between the two VKC-2 tasks (lunch, breakfast) was tested using the Spearman Brown formula (r). Coefficients > 0.70 were interpreted as evidence for strong internal consistency[81].

## Results

### Participant Characteristics

237 participants were recruited from June of 2021 to June of 2025 for studies on everyday function. One participant with mild dementia refused to complete the study tasks; thus, the final analytic sample included 236 participants, of which 172 participants were classified as healthy cognition, 48 as MCI, and 16 as mild dementia. On average, participants were 72 years old, completed 15 years of education, included 66.1% (156/236) women and nearly equal numbers of people who identified as Black (44.9%; n=106/236) versus White (47.9%; n=113/236) race. Demographic characteristics of the groups are reported in Table 2. The groups differed in age and education. Post hoc comparisons did not reach statistical significance, but the healthy cognition and MCI groups differed most on age (p = .056) and the healthy cognition and dementia groups differed the most on education (p = .051). There were no group differences in estimated IQ or in the distributions of sex, Black/African American vs. White race, or ethnicity.

**Table 2.**
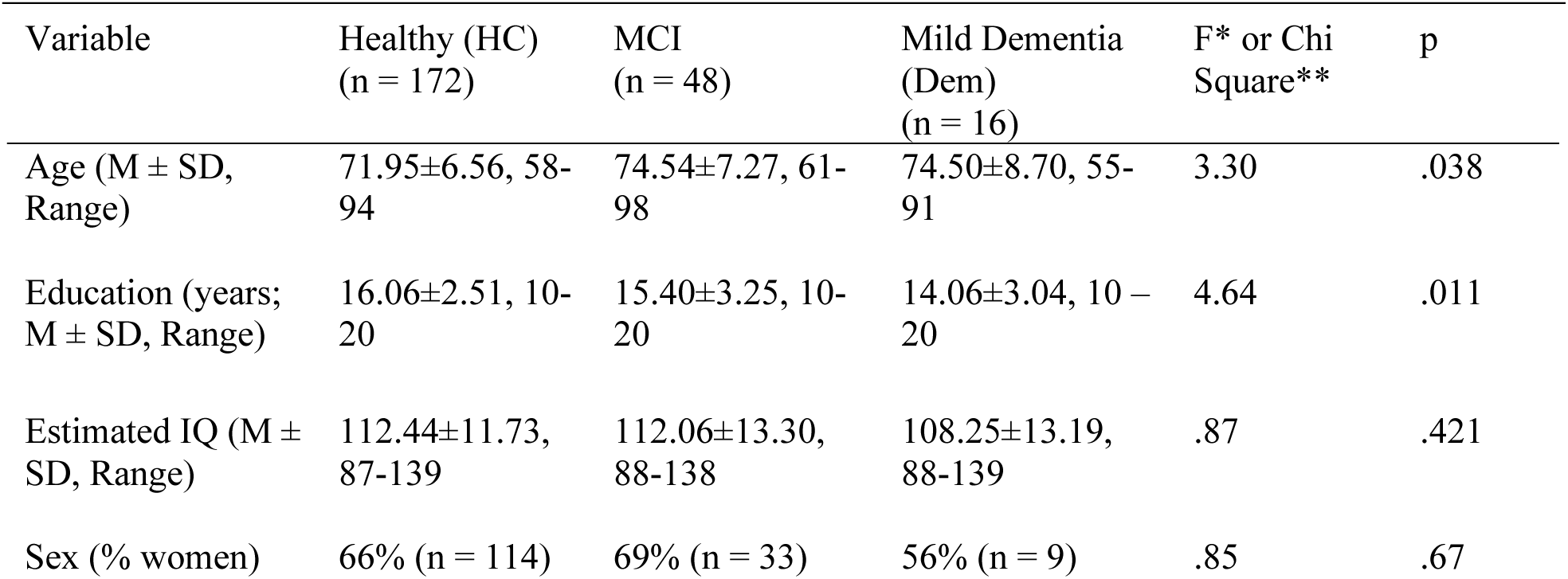
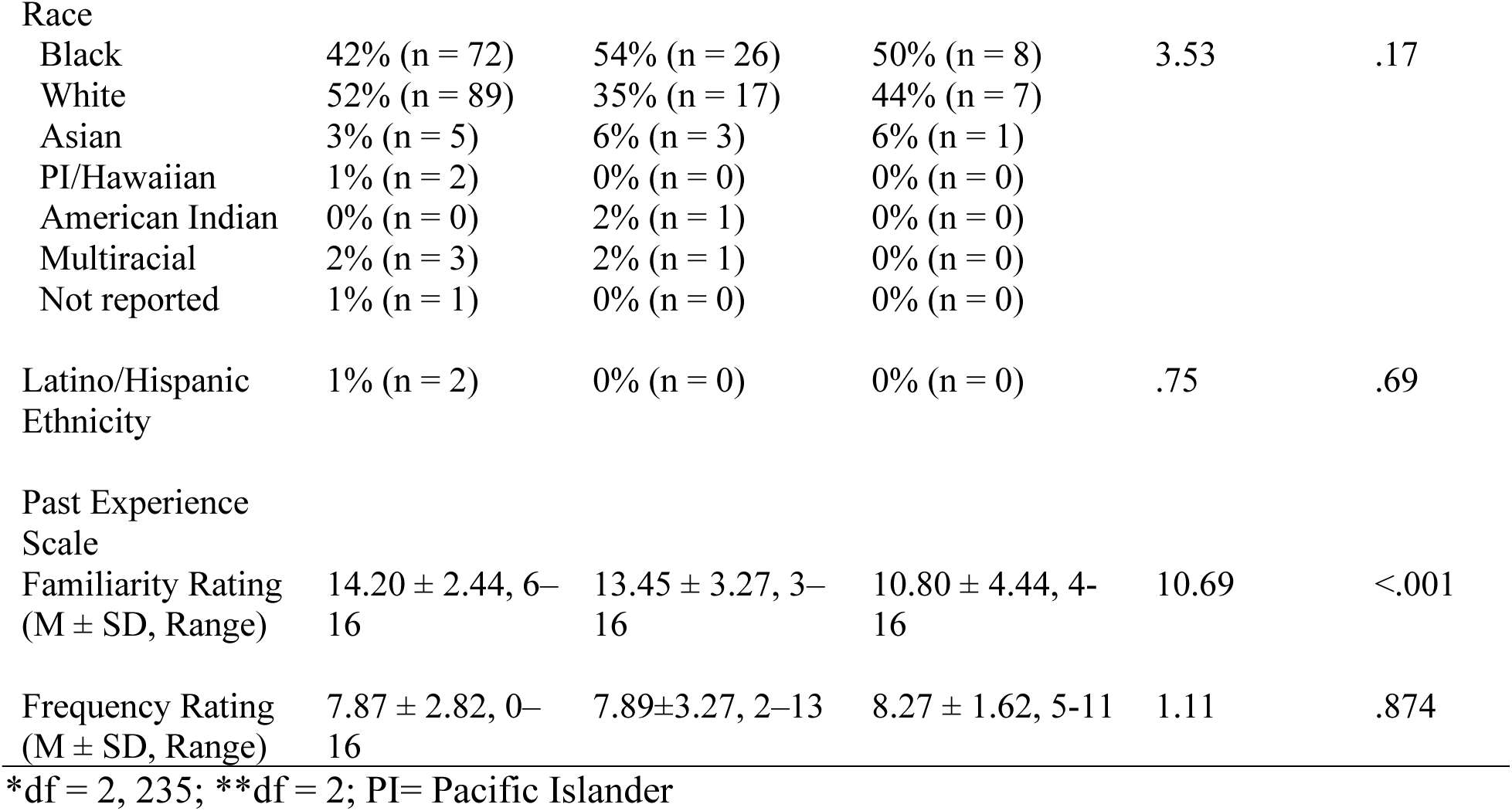
Demographic and descriptive characteristics by group.

Results from the Past Experience Scale showed Task Familiarity ratings were generally high, indicating that on average the breakfast/lunch tasks were “pretty” to “very” familiar. The groups differed on the Familiarity Rating, with post-hoc tests indicating that the healthy group reported significantly higher task familiarity than the dementia group (p =.027); all other group comparisons were not statistically significant. According to the Frequency Ratings, participants reported that on average they perform the VKC-2 tasks about once per month over the past 5-10 years. Frequency Ratings did not differ across the groups.

Demographic characteristics of participants who returned for session 2 and who were included in the retest reliability analysis are reported in Supplemental Materials. Compared to participants who did not return, the group of returning participants had completed significantly more years of education, obtained higher estimated IQ scores, and included a greater proportion of White participants.

### Informant Characteristics

219 informants participated in the study. On average, informants were 63.97 years old (SD = 13.99, range 20 – 90) and completed 15.73 years of education (SD = 2.43, range 10 – 21). Informants included spouses (43.4%; n=95/219), children (27.4%; n=60/219), friends (18.7%; n=41/219) or other family members (10.5%; n=23/219).

### Correlations among VKC-2 Scores

Average scores on the VKC-2 and their bivariate correlations are shown in Table 3. On average, participants took about 87 seconds to complete the Movement Familiarization Task (as reflected by the digital dexterity score). On the VKC-2 Test, average completion time was 197 seconds with an average of 68 screen interactions. On average, participants spent approximately 48% of the completion time off the screen. As stated, the distractor interactions score was dichotomized because there were few instances of distractor interactions, with only 8.9% (21/237) of participants interacting with distractor objects. Bivariate correlations indicate significant associations among all scores. The relation between VKC-2 time and touches was the strongest and reflected nearly overlapping scores, as more touches were associated with longer completion times.

**Table 3.** VKC-2 scores and correlation coefficients in the full sample (N =236).

|  | Digital Dexterity | Time | Touches | %Off Screen |
| --- | --- | --- | --- | --- |
| Time | .67* | — |  |  |
| Touches | .51* | .83* | — |  |

|  | Digital<br>Dexterity | Time | Touches | %Off Screen |
| --- | --- | --- | --- | --- |
| %Off Screen | .49* | .42* | .34* | — |
| Distractor Interactions | .34* | .39* | .42* | .26* |
| Mean | 86.76 | 197.47 | 67.82 | .48 |
| SD | 27.47 | 111.57 | 50.59 | .09 |

\*p <.001 (2 tailed)

### Construct Validity

The construct validity of the VKC-2 automated scores was evaluated by assessing differences among groups with known differences in functional abilities: healthy, MCI, and mild dementia. As shown in Figure 3, average scores were consistently worse for the dementia group, but statistical analyses focused on differences between the healthy and MCI groups, because of the relatively few dementia participants. ANCOVA results comparing healthy vs. MCI participants are reported in Table 4 and showed significant group differences in all measures after controlling for age. After controlling for the digital dexterity score and age (see bottom of Table 4), the difference in time was no longer significant, suggesting the difference in time could be explained by low-level visuomotor skill differences between the MCI and healthy groups. After controlling for the digital dexterity score, the differences in touches and %off-screen remained statistically significant, suggesting between group differences in these scores could not be explained by differences in basic visuomotor skills.

**Figure 3.**
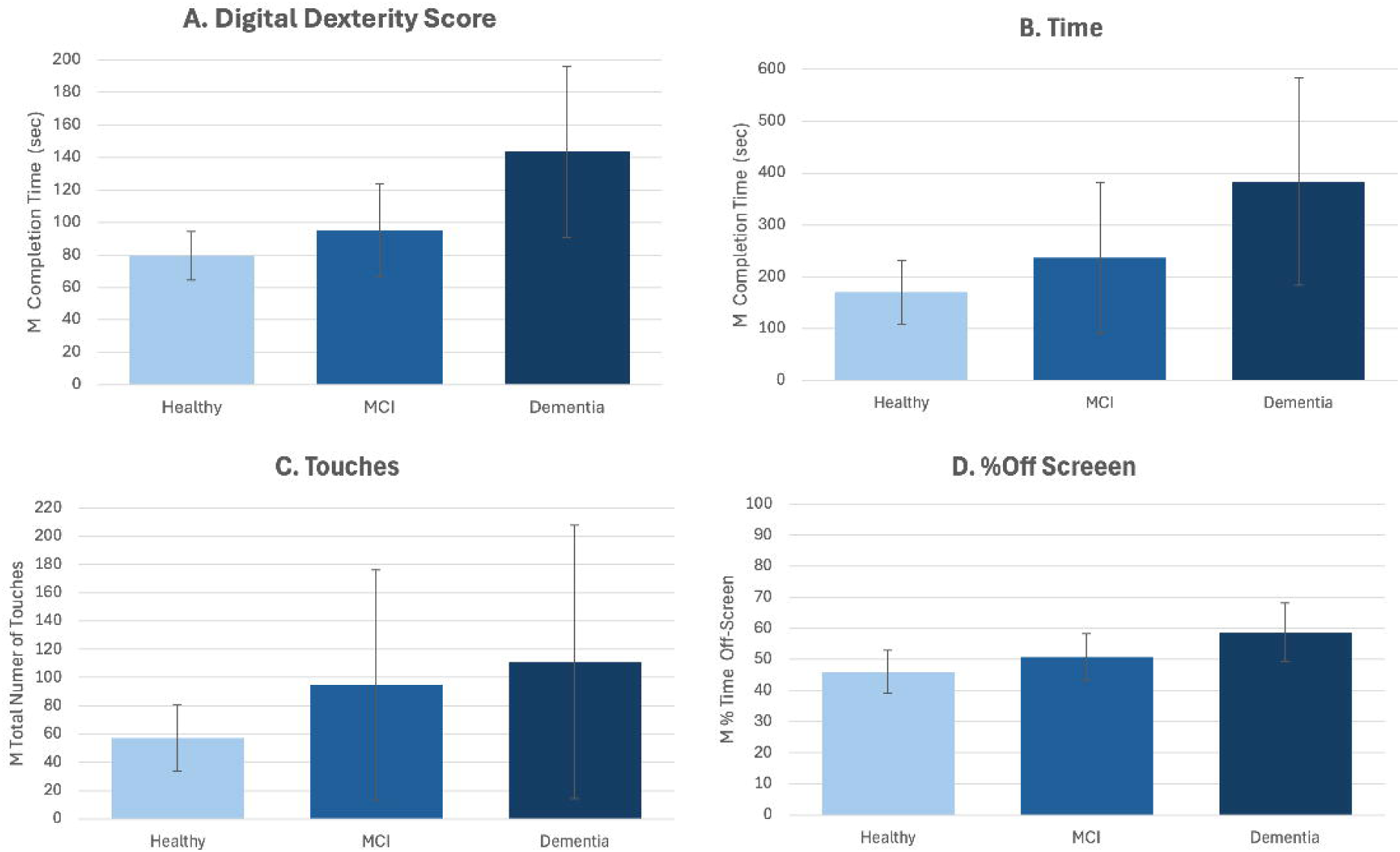
(Unadjusted VKC-2 mean scores by group)

**Table 4.** ANCOVA results comparing healthy cognition (n = 172) vs. MCI (n = 48) groups on all VKC-2 automated scores.

| Controlling for Age (df = 2, 219) |  |  |  |  |
| --- | --- | --- | --- | --- |
| VKC-2 Score | F | p | $\eta^2$ (partial eta<sup>2</sup>) | Effect Size |
| Digital Dexterity | 20.68 | < .001 | 0.087 | medium- large |
| Time | 17.54 | < .001 | 0.075 | medium |
| Touches | 23.35 | < .001 | 0.097 | large |
| %Off-Screen | 12.71 | < .001 | 0.055 | medium |

Controlling for Age and Digital Dexterity (df = 3, 219)
| VKC-2 Score | F | p | $\eta^2$ (partial eta<sup>2</sup>) | Effect Size |
| --- | --- | --- | --- | --- |
| Time | 3.47 | .064 | .016 | small |
| Touches* | 8.60 | .004 | .038 | small - medium |
| %Off-Screen* | 6.42 | .012 | .029 | small - medium |

Effect sizes are interpreted as: small = .01, medium = .06, large = .14

The distributions of the distractor interaction score across the three groups (not reported in Figure 3) showed a higher percentage of participants interacted with distractors in the groups with cognitive impairment (dementia = 31.3%; n =5/16; MCI = 16.7%; n =8/48; heathy = 4.7%, n=5/114). The difference between MCI and the healthy groups in distractor interactions was statistically significant (χ² (1) =8.03, p = .005; φ = .191, small-to-medium effect size).

### Classification Analyses

Classification analyses for distinguishing participants with healthy cognition from participants with cognitive impairment (MCI + mild dementia group combined) are reported in Table 5. All predictors showed statistically significant areas under the curves (AUC), indicating that they were better than chance at predicting impaired group status. Time was the strongest predictor, as indicated by the highest AUC and sensitivity, making it useful to maximize identification of impaired participants for early detection. %off-screen showed the highest specificity suggesting that %off-screen may be more useful in ruling out people with healthy cognition for diagnostic confirmation. Analyses for distinguishing participants with healthy cognition versus MCI demonstrated similar AUC (.68. -.71), cut scores, and patterns of sensitivity specificity and are reported in Supplemental Materials.

**Table 5.** Area Under the Curve (AUC) values, optimal cutoffs, and specificity/sensitivity for predicting cognitive impairment from VKC-2 scores (N =236).

| VKC-2 Score | AUC | 95% CI<br>(Lower–Upper) | SE | p-value | Optimal<br>Cutoff | Sensitivity | Specificity |
| --- | --- | --- | --- | --- | --- | --- | --- |
| Digital Dexterity | .73 | .65-.81 | .41 | < .001 | 87.12 | .67 | .77 |
| Time | .75 | .68-.82 | .04 | < .001 | 163.47 | .82 | .58 |
| Touches | .70 | .62-.78 | .04 | < .001 | 65.5 | .59 | .78 |
| %Off-Screen | .71 | .64-.79 | .04 | < .001 | .53 | .48 | .87 |

### Convergent Validity against Real Kitchen Scores

Bivariate correlations between VKC-scores and scores on the Real Kitchen are reported in Table 6. Coefficients with Real Kitchen Completion Time, Accomplishment and Total Errors were consistently significant and moderate to strong, indicating convergent validity of the VKC-2 and scores against real versions of the VKC-2 tasks (i.e., Real Kitchen). Relations between VKC-2 measures and motor errors on the Real Kitchen were weak and not consistently significant, suggesting that the VKC-2 scores may correspond more strongly with the cognitive aspects of the Real Kitchen performance rather than visuomotor errors made with the real tasks. Spearman rank order correlations showed the same pattern of results and are reported in Supplemental Materials.

**Table 6.**
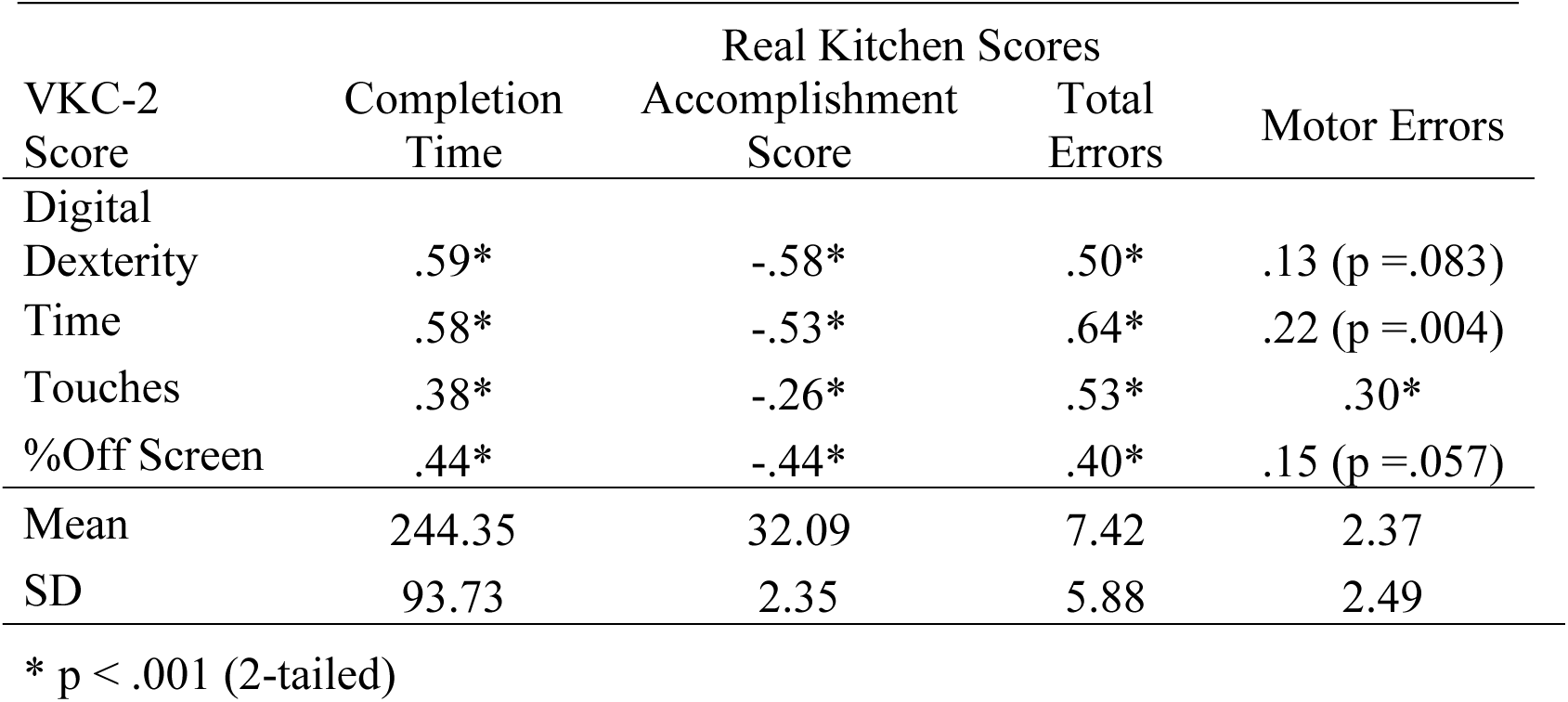
Correlation coefficients (and p-values) between VKC-2 scores and Real Kitchen scores (n = 201).

### Convergent Validity against Conventional Cognitive Tests

Bivariate correlations between VKC-2 scores and demographically adjusted cognitive test scores are reported in Table 7. Coefficients were statistically significant and indicated that that participants with better cognitive test scores completed the VKC-2 tasks more quickly and efficiently, supporting the convergent validity of the VKC-2 scores. Spearman rank order correlations showed the same pattern of results and are reported in Supplemental Materials.

**Table 7.**
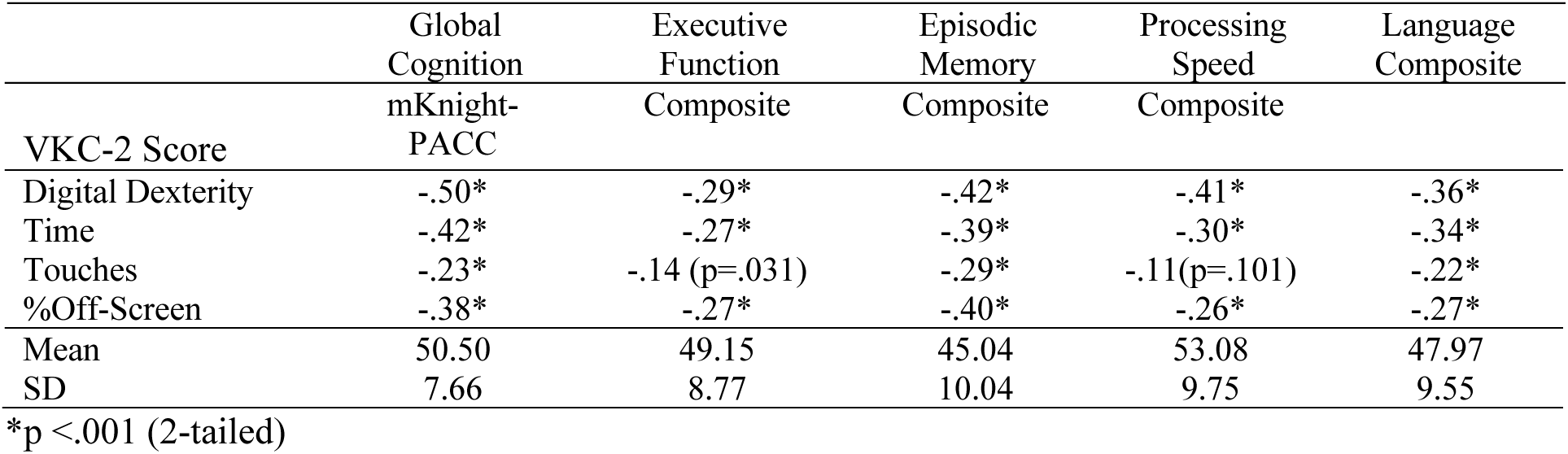
Correlation coefficients (and p-values) between VKC-2 scores and cognitive test scores (N = 236)

### Convergent Validity against Self/Informant Questionnaires of Everyday Function

Table 8 shows relations between VKC-2 scores and questionnaires regarding everyday function completed by participants and informants. Results from participant questionnaires showed that relations between the VKC-2 scores and the IADL-C and FAQ, which assess current functional abilities were statistically significant and in the expected direction. That is, participants who reported greater current functional difficulties (IADL-C, FAQ) also performed the VKC-2 less quickly and efficiently. The relation between the VKC-2 scores and participant’s report of functional decline (ECog) was not significant. By contrast, informant reports of both current functional difficulties (IADL-C, FAQ) and functional decline (ECog) were significantly associated with lower VKC-2 scores. Overall, correlations between the VKC-2 and participant/informant questionnaires support the validity of the VKC-2. Spearman rank order correlations showed a similar pattern of results and are reported in Supplemental Materials.

**Table 8.**
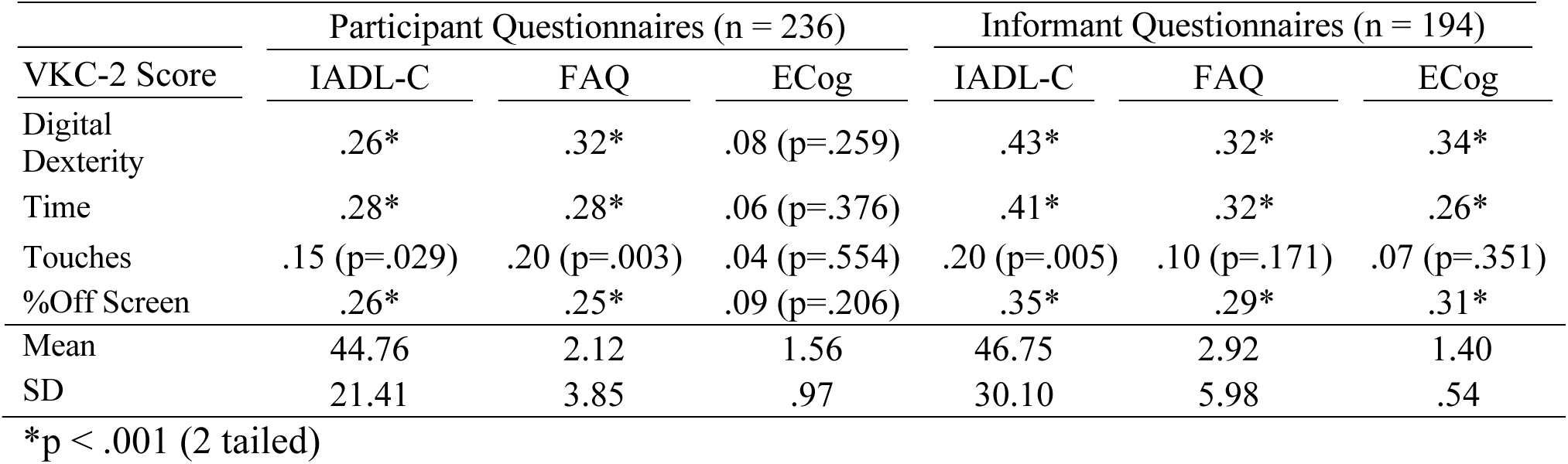
Correlation coefficients (and p-values) between VKC-2 scores and questionnaires.

| VKC-2 Score | Participant Questionnaires (n = 236) |  |  | Informant Questionnaires (n = 194) |  |  |
| --- | --- | --- | --- | --- | --- | --- |
|  | IADL-C | FAQ | ECog | IADL-C | FAQ | ECog |
| Digital Dexterity | .26* | .32* | .08 (p=.259) | .43* | .32* | .34* |
| Time | .28* | .28* | .06 (p=.376) | .41* | .32* | .26* |
| Touches | .15 (p=.029) | .20 (p=.003) | .04 (p=.554) | .20 (p=.005) | .10 (p=.171) | .07 (p=.351) |
| %Off Screen | .26* | .25* | .09 (p=.206) | .35* | .29* | .31* |
| Mean | 44.76 | 2.12 | 1.56 | 46.75 | 2.92 | 1.40 |
| SD | 21.41 | 3.85 | .97 | 30.10 | 5.98 | .54 |

\*p < .001 (2 tailed)

### Retest Reliability

Intraclass correlation coefficients (ICC) results are reported in Table 9 and indicated good to excellent reliability for the VKC-2 automated scores. Cohen’s kappa to assess agreement between distractor interaction scores at Time 1 and Time 2 showed fair agreement, κ = 0.27, p < .001, indicating limited but statistically significant consistency over time. When ICC were re-run including only participants who reported no change in their cognitive status from session 1 (86.0%; 123/143), results yielded comparable or slightly improved coefficients/reliability relative to the full sample (see Supplemental Materials).

**Table 9.**
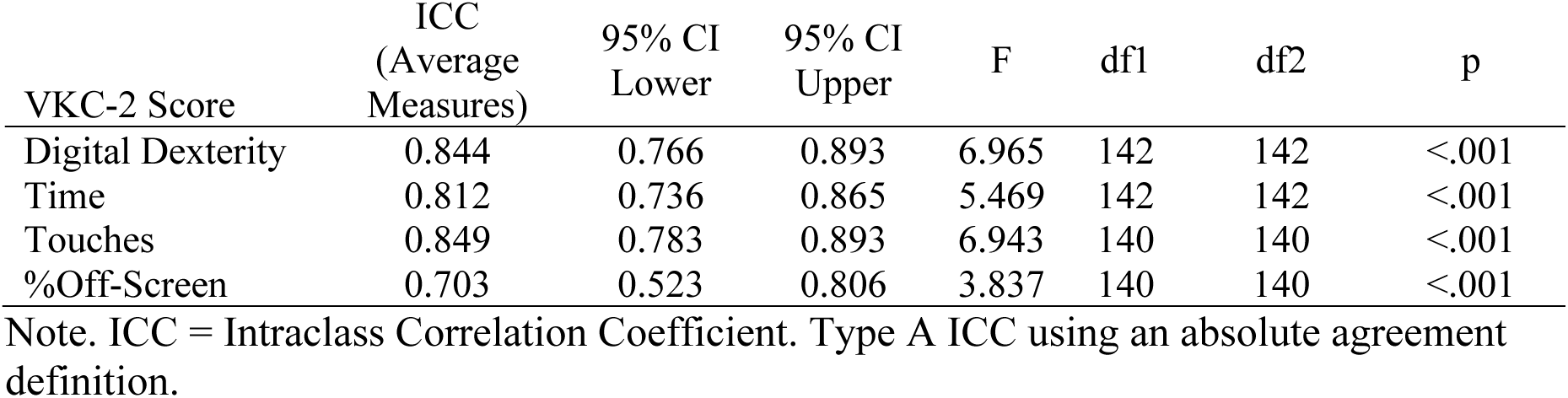
Intraclass correlation coefficients for VKC-2 scores over time (n = 143)

| VKC-2 Score | ICC<br>(Average<br>Measures) | 95% CI<br>Lower | 95% CI<br>Upper | F | df1 | df2 | p |
| --- | --- | --- | --- | --- | --- | --- | --- |
| Digital Dexterity | 0.844 | 0.766 | 0.893 | 6.965 | 142 | 142 | <.001 |
| Time | 0.812 | 0.736 | 0.865 | 5.469 | 142 | 142 | <.001 |
| Touches | 0.849 | 0.783 | 0.893 | 6.943 | 140 | 140 | <.001 |
| %Off-Screen | 0.703 | 0.523 | 0.806 | 3.837 | 140 | 140 | <.001 |

Note. ICC = Intraclass Correlation Coefficient. Type A ICC using an absolute agreement definition.

### Internal Reliability

Internal consistency between the VKC-2 breakfast and lunch tasks at Time 1 was evaluated using Spearman-Brown coefficients in the full sample (N = 236). Results showed acceptable to good internal consistency for all scores (time .81; touches .81; %off-screen .77).

## Discussion

Results of the current study support the validity and reliability of the VKC-2 automated scores as measures of everyday function in older adults. As predicted, VKC-2 automated scores differed significantly between groups known to differ in functional ability level (healthy vs. MCI vs. mild dementia) supporting the construct validity of the VKC-2. Convergent validity was supported by the significant correlations between VKC-2 automated scores and performance on real versions of the VKC-2 tasks (Real Kitchen), scores on conventional cognitive tests, and self/informant questionnaires regarding everyday functioning. In terms of reliability, ICC indicated good to excellent retest reliability for the VKC-2 automated scores. There was also good internal reliability of the VKC-2 tasks (breakfast, lunch). Additionally, participants reported that the tasks included in the VKC-2 were highly familiar (Past Experience Questionnaire). Our findings suggest that the VKC-2 automated scores have great promise for addressing important gaps in functional assessment for multiple uses and contexts, including screening older adults at risk for decline in meaningful, functional activities in primary care and as a function endpoint in clinical trials of AD/ADRD treatments.

To our knowledge this is the study to show significant differences between older adults with healthy cognition vs. MCI on the VKC-2 automated scores. These results are consistent with the findings of a previous version of the Virtual Kitchen that found significant differences between participants with dementia (AD) versus healthy controls on completion time and error scores[57] as well as a prior study that demonstrated significant differences between healthy young adults versus healthy older adults[39]. Our group and others have demonstrated significant differences between participants with MCI and healthy cognition on performance-based tasks with real objects[26,32,33,45]. McKniff and colleagues[35] also demonstrated significant differences in completion time, errors, and efficiency between participants with healthy cognition and MCI on the real versions of the VKC-2 tasks (Real Kitchen). Thus, the current results are in line with past pilot studies of earlier versions of the Virtual Kitchen and mirror findings from performance-based tests with real objects. Taken together, the results support the utility of the VKC-2 to identify everyday task difficulties without need for videorecording or trained coders, offering a major advantage over traditional performance-based tests for a highly efficient, scalable, and sensitive measure of everyday functioning.

To our knowledge our results also are the first to show VKC-2 differences between groups with known functional differences after adjusting for digital visuomotor dexterity, ensuring that the differences between MCI and healthy participants cannot be explained simply as a difference in digital visuomotor skills or accuracy in touchscreen interactions. Even after controlling for digital dexterity, healthy controls and individuals with MCI differed significantly on measures of task efficiency (touches, %off-screen). Prior work on an earlier version of the Virtual Kitchen has shown that some time off-screen is spent making micro-errors toward the screen without screen contact[39]. A comparable measure derived from a digital clock drawing test has shown that compared to healthy controls, older adults with memory impairment spent a significantly greater proportion of their clock drawing time off-screen (i.e., thinking or planning)[83]. Thus, the %off-screen score captures meaningful erroneous and inefficient behaviors that are not measured through direct contact with the screen.

The difference in VKC-2 completion time between participants with MCI vs. healthy cognition did not remain significant after controlling for digital dexterity, suggesting that VKC-2 completion time is influenced by visuomotor skill. VKC-2 time scores may offer useful information regarding real world functional abilities, as mild upper motor dexterity difficulties contribute to functional difficulties in people with MCI[20] and mild upper and lower limb difficulties are significantly associated with cognitive difficulties in older adults[21,22,84]. Indeed, the VKC-2 digital dexterity score was associated with a measure of global cognitive abilities (mKnight-PACC) that is sensitive to preclinical AD. Thus, mild motor difficulties may be important early indicators of AD/ADRD risk that could be missed by conventional cognitive tests. Future work, including longitudinal follow up is needed to identify the best combination of VKC-2 scores to maximize early detection of functional difficulties and risk.

Correlation analyses also supported the convergent validity of the VKC-2 automated scores. VKC-2 scores were strongly and consistently associated with measures of conceptual accuracy and performance efficiency on the real versions of the VKC-2 tasks (Real Kitchen). Past studies examining virtual performance-based assessments of real-world tasks have reported similar results. For example, Allain and colleagues[57] showed positive relations between a real-world coffee making task and a non-immersive virtual coffee-making task in participants with dementia and healthy cognition, and a case report of a 91-year-old woman showed similar performance patterns in a real and virtual breakfast task[85]. The VKC-2 was designed to simulate real tasks and assess the same cognitive operations recruited during the performance of real everyday tasks—object selection, sequencing of steps, goal maintenance, and performance monitoring[72,86,87]. The strong significant associations between the VKC-2 and Real Kitchen scores support the conceptualization of the VKC-2 as a measure of the same cognitive operations required to complete the real-world tasks.

One difference between the Real Kitchen and the VKC-2 is that VKC-2 scores were not significantly associated with motor errors on the Real Kitchen (i.e., instances of imprecision that are not conceptualized to reflect cognitive failures in task performance). This is consistent with previous findings with an earlier version of the VKC[39] and suggests that the visuomotor demands of the Real Kitchen are not captured with the VKC-2 scores. Motor errors on the Real Kitchen have not been extensively studied and have been considered to have little theoretical interest in prior work focused on the cognitive aspects of everyday function[27,72,74,86]. Additional research is needed to determine whether motor errors on the Real Kitchen have predictive validity and whether the VKC-2 may be lacking because it does not measure the same visuomotor abilities as the Real Kitchen.

Correlation analyses with conventional clinical measures, including cognitive tests and self/informant questionnaires of everyday function showed further support for the validity of the VKC-2 automated measures. All VKC-2 scores were moderately associated with demographically adjusted cognitive composite scores consistent with correlation coefficients reported for other performance-based tests and questionnaires of everyday function[88,89]. Relations were observed with the mKnight-PACC, a cognitive composite score shown to be sensitive to preclinical AD, and with domain-specific composite scores[63]. Associations with domain scores of episodic memory and language, which do measure motor skills or processing speed, are particularly noteworthy and offer further support that the automated VKC-2 scores reflect cognitive abilities beyond visuomotor skills consistent with results from an earlier version of the Virtual Kitchen[39]. In a recent study of young adults, VKC automated scores were associated with highly sensitive measures of executive attention, reflecting the sensitivity of VKC-2 scores for measuring cognitive abilities even among cognitively healthy younger people[52]. The significant association of the VKC-2 measures with conventional cognitive tests add to the growing interest of digital cognitive assessments with sensitive and novel measures that reflect cognitive abilities.

Significant correlations between VKC-2 scores and questionnaires of functional abilities also support the validity of the VKC-2 scores as measures that are meaningful regarding real world, everyday function. Interestingly, correlations were stronger and more consistent with informant report, particularly for the questionnaire assessing cognitive/functional decline (ECog[13]). Thus, although the VKC-2 scores may be used to estimate everyday functioning in real life when informants are not available or not reliable, we acknowledge that the VKC-2 and questionnaires measure slightly different constructs pertaining to everyday function. The VKC-2 is a measure of everyday functional capacity in a controlled environment, making it ideal for between participant comparisons and tracking change over time. Questionnaires assess functioning in a highly unconstrained context in which task demands, motivation, economic resources, social support, and a wide range of other factors are also at play. Thus, ideally, particularly for clinical purposes, the VKC-2 would be used in conjunction with questionnaires for a comprehensive assessment of everyday function across contexts.

To our knowledge ours is the first study to evaluate the reliability of any version of the Virtual Kitchen. Retest reliability estimates (ICC) showed automated VKC-2 scores, except for distractor interactions, which occurred very infrequently, were stable over the span of 4 -6 weeks. ICC were even stronger after excluding participants who reported a notable change in their cognitive abilities. Strong test-retest reliability is crucial to evaluate meaningful change over time and for use in clinical trials. VKC-2 tasks (breakfast, lunch) also demonstrated strong internal consistency, supporting the coherence of the combined, total VKC-2 scores.

Several study strengths and weaknesses are worth noting. Among the strengths is the sample size and the inclusion of many participants (42%) who reported Black or African American race. The racial diversity of the sample addresses a critical gap in cognitive assessment research and enhances the generalizability of our findings to clinical populations across the US. The portability and automated scoring and standardized administration protocol of the VKC-2 is a clear strength over current function measures and existing regulatory-approved outcome measures for clinical trials, which require specialized training, lengthy administration times, and often unavailable informants. The efficiency of the VKC-2 over conventional cognitive test batteries also offers advantages for busy clinical settings where comprehensive neuropsychological batteries are impractical.

Study limitations also warrant consideration. First, while our sample included substantial racial diversity, the predominance of highly educated participants (mean 15.7 years) may limit generalizability to populations with lower educational attainment. Additionally, our sample was majority women (66%) and fell short in representation of races other than Black/African American or White and Latino/Hispanic ethnicity. The current community-based sample also included mostly older adults with healthy cognition, with only 27% of participants meeting criteria for impaired cognition. The imbalance in the size of subgroups with healthy versus impaired cognition limited power for between-group statistics and AUC/classification analyses. Thus, additional work is needed to replicate these findings in samples with larger groups of people with MCI or mild dementia. Second, the virtual task environment, while ecologically valid, may not capture all aspects of real-world functional demands such as physical fatigue, environmental distractions, or competing task demands. Third, our cross-sectional design limited conclusions about the ability of the VKC-2 to detect meaningful change over time or predict clinical outcomes (predictive validity). Finally, direct validation against regulatory outcome measures is needed before considering the VKC-2 as an alternative endpoint in clinical trials.

Important future directions for research on the VKC-2 includes longitudinal study to determine the predictive validity of the VKC-2 scores. It would be important to know whether the VKC-2 outperforms conventional measures in identifying individuals who experience cognitive and functional decline. However, even if the VKC-2 is only just as good as conventional cognitive measures or questionnaires, it has important advantages over conventional measures because of its efficiency and because it does not depend on a reliable informant. Another important future direction is validation against biomarkers of neurodegenerative disease. Holmqvist and colleagues[48] demonstrated strong correlations between VKC-2 scores and MRI-derived measure of cerebral white matter hyperintensities, a biomarker of cerebral small vessel disease associated with brain aging and neurodegenerative disease. Future studies investigating associations between VKC-2 scores and other biomarkers, including AD-specific PET and blood biomarkers are in progress. Finally, automated VKC-2 scores reflecting task accomplishment are under development and will expand the utility of the VKC-2 as a tool to deepen our understanding of difficulties with everyday task performance through detailed characterization of performance patterns[86]. Future implementation research should examine potential barriers to VKC-2 adoption, including technology comfort levels across diverse older adult populations and integration with existing clinical workflows.

In conclusion, there is a growing interest in the development of digital assessments of cognition including digitized versions of traditional tests of cognition, smart-phone and tablet based cognitive assessment, and virtual reality[82]. Digital, performance-based assessments of everyday tasks, like the VKC-2, extend this trend to meaningful measures of everyday function. The VKC was designed to address weaknesses of conventional functional measures function though objective, standardized, highly efficient assessment without informant report. The VKC-2 requires approximately 15 to 20 minutes to administer, is appropriate for the full spectrum of cognitive aging, from healthy aging to mild dementia, and includes tasks (lunch, breakfast) that have been extensively studied and shown to be highly familiar to older adults[27,28,32,47,91]. The VKC-2 may be administered on a portable, laptop administration without need for objects or other supplies, including a VR headset, precluding limitations associated with cyber sickness and confusion. The touch screen interface is more natural than mouse or joystick for older adults[92]. Finally, the VKC-2 yields sensitive and detailed analysis of performance, including time to completion and measures of performance efficiency derived from the touch screen, eliminating the need for video recording and human coders. The older adult participants in this study were capable of using the touch-screen interface, understood the directions, did not require extensive training, and performed the tasks consistent with expectations based on data from the Real Kitchen[35]. The VKC-2 shows great promise as an ecologically valid and scalable tool for capturing everyday functional capabilities in people with healthy cognition, MCI, and mild dementia across settings, including large, longitudinal studies, health clinics, and clinical trials.

## Data Availability

All data produced in the present study are available upon reasonable request to the authors.

## Acknowledgments

The authors acknowledge Anna Callahan, Julina Hossfeld, Nicole Lloyd, Julia Manganti, Shrey Patel, Emma Pinsky, and Yuki Tsuchiya for their contributions to the Virtual Kitchen Challenge Project, which included participant recruitment, assistance with data collection, and database management. The authors also are grateful to Ms. Sherry Hill, director of the Community Engagement Fitness Network for her help in community outreach and participant recruitment.

## Data Availability

The dataset analyzed for this study will be available in the Virtual Kitchen Challenge Repository at DOI 10.17605/OSF.IO/MJXWE. In the interim, data requests may be sent to the corresponding author.

## Important Note on Generative AI

Generative AI was not used in the creation of this manuscript.

## Authors’ Contributions

MK, MM, SMS., MBT., KH, SH, RM, KH, RC, MR, GV, and MDS contributed to data collection and implementation. MK, MM, and TG contributed to writing (original draft preparation). MK, MM, TG, MDS, SMS, MBT, REM, KH and KH contributed to writing (review and editing). TG and TY contributed to methodology and conceptualization. TG and DAGD contributed to formal analyses. TG contributed to project administration and funding acquisition. All authors read and agreed to the version of the manuscript intended for publication.

## Abbreviations

(AD/ADRD): Alzheimer’s disease related disorders
(MCI): Mild cognitive impairment
(VKC-2): Virtual Kitchen Challenge – Version 2
(VKC): Virtual Kitchen Challenge
(NAT): Naturalistic Action Test
(VR): Virtual Reality
(mKnight-PACC): Knight-Preclinical Alzheimer Cognitive Composite
(ECog): Everyday Cognition questionnaire
(FAQ): Functional Activities Questionnaire
(IADL-C): Instrumental Activities of Daily Living—Compensation questionnaire
(ICC): Intraclass Correlation Coefficients
(AUC): Area Under the Curve

