## Supplemental Materials for "The Virtual Kitchen Challenge–Version 2: Validation of a Digital Assessment of Everyday Function in Older Adults"

**Supplemental Materials for The Virtual Kitchen Challenge–Version 2: A Valid, Reliable, and Efficient Digital Tool for Measuring Everyday Function in Older Adults**

**Procedures used to classify participants as having healthy cognition vs. MCI vs. mild dementia.**

Participants were classified as having healthy cognition, MCI, or mild dementia using demographically adjusted (age, education, sex, and race) T scores from the cognitive tests and according to Jak/Bondi neuropsychological actuarial classification criteria[1,2].

*Healthy Cognition Criteria*: meet general criteria (described above); no self or informant report of significant cognitive decline (i.e., never sought clinical exam for cognitive concerns); no self or informant report of dementia/MCI diagnosis; no significant functional difficulties noted on self- or informant report (Functional Activities Questionnaire; FAQ < 9); no two demographically adjusted T scores within at least one cognitive domain below the normative mean (T score = 40) nor one T score in each of the four cognitive domains (episodic memory, working/memory, executive functioning, language, and processing speed) on cognitive tests administered at session 1.

*MCI Criteria*: prior diagnosis of MCI (any subtype); confirmation of no significant functional difficulties reported on informant report (FAQ < 9); confirmation of mild cognitive impairment based on tests administered during Session 1 [i.e., demographically adjusted cognitive test scores that are 1 SD worse than normative mean on both measures within at least one cognitive domain (episodic memory, working memory/executive functioning, language, processing speed) or one score 1 SD worse than the normative mean in each of the four cognitive domains sampled].

*Mild (all-cause) Dementia Criteria:* diagnosis of mild dementia (any subtype); confirmation of significant functional difficulties reported on informant report (FAQ > 9 at session 1); confirmation of cognitive impairment on demographically adjusted cognitive test scores (i.e., approximately 1.5 SD worse than normative mean in at least one cognitive domain- episodic memory, working memory/executive functioning, language, processing speed) at session 1.

**Human Coding Procedures for the Real Kitchen**

All videos were scored by two coders blind to participant characteristics. See McKniff and colleagues[1] for details on inter-rater reliability and scoring guidelines. The following scores were obtained following review and consensus between the two separate coders:

**Completion Time** -The amount of time (in seconds) the participant took to complete the breakfast and the lunch task. Time elapsed from first movement toward object to when they pressed the quit button. A higher completion time suggests worse performance.

**Accomplishment** - The number of steps participants accurately completed. This score includes the sum of the accomplishment steps for the breakfast and lunch tasks (out of 33 total possible steps). A higher score is indicative of better performance (more steps accomplished).

**Total Errors** - Sum of all overt commission errors and micro-errors. Overt commission errors include instances when a step is performed inaccurately (e.g., wrong sequence or with the wrong object) and instances when an additional step is performed (e.g., action addition, perseveration). Micro-errors include any subtle inexact action that might not reach the level of an error but is still not accurate and is inefficient. Micro-errors include misreaching actions that are pre-emptively corrected, including instances when the participant changes trajectory of their reaching movement. Micro-errors also include instances when participants touch objects that they do not immediately use and when an object is moved purposelessly or without using it. A total error score was computed for errors made during the breakfast and lunch tasks; a higher score is indicative of worse performance.

**Motor Errors** – Instances when a correct step is performed with some visuomotor imprecision, such as spilling coffee grinds on the table when adding coffee to the mug or dropping the thermos lid when sealing the thermos as part of the lunch task. A higher motor error score suggests more visuomotor difficulties.

Supplementary Table 1. Demographic characteristics of participants who returned for the second session (n = 143) versus participants who did not return (n =93)

| Variable | Participants who Returned for Visit 2 (n = 143) | Participants who did NOT Return for Visit 2 (n = 93) | t* or Chi Square | p |
|---|---|---|---|---|
| Age (M ± SD, Range) | 73.29±6.48, 61-98 | 71.58±7.33, 55-86 | -1.88 | .061 |
| Education (years; M ± SD, Range) | 16.11±2.74, 10-20 | 15.37±2.69, 10-20 | -2.06 | .041 |
| Estimated IQ | 114.34±11.79, 87-139 | 108.63±12.0, 89-138 | -3.60 | <.001 |
| Sex (% women) | 62.2% (n = 89) | 73.1% (n =68) | 3.00 (df=1) | .083 |
| Race | | | 16.88 (df =1) | <.001 |
| Black | 33.6% (n = 48) | 62.4% (n =58) | | |
| White | 57.3% (n = 82) | 33.3% (n=31) | | |
| Asian | 4.9% (n = 7) | 2.2% (n=2) | | |
| PI/Hawaiian | 1.4% (n = 2) | 0% (n = 0) | | |
| Native American | 0% (n = 0) | 1% (n = 1) | | |
| Multiracial | 1.4% (n = 2) | 1% (n = 1) | | |
| Not reported | 1% (n = 1) | 0% (n = 0) | | |
| Latino/Hispanic Ethnicity | 1% (n = 1) | 1% (n = 1) | .10 (df=1) | .758 |
| Clinical Classification | | | .40 (df=2) | .817 |
| Healthy Cognition | 74.1% (n=106) | 71.7% (n=66) | | |
| MCI | 20.3% (n=29) | 20.7% (n=19) | | |
| Dementia | 5.6% (n=8) | 7.6% (n=7) | | |

PI = Pacific Islander; *df =234

**Classification Analyses for Predicting MCI versus Healthy Cognition**

All predictors showed statistically significant AUCs, indicating that they were better than chance at predicting MCI group status. Time was the strongest predictor, as indicated by the highest AUC and sensitivity, making it useful to maximize identification of MCI participants for early detection. Touches and the digital dexterity score show better specificity indicating that these scores may be more useful in ruling out people with healthy cognition for diagnostic confirmation.

Supplementary Table 2. Area Under the Curve (AUC) values for predicting MCI group membership.

| VKC-2 Score | AUC | 95% CI (Lower–Upper) | SE | p-value | Optimal Cutoff | Sensitivity | Specificity |
|---|---|---|---|---|---|---|---|
| Digital Dexterity Score | .68 | .60–.78 | .046 | < .001 | 87.12 | 0.60 | 0.77 |
| Time | .71 | .63–.79 | .042 | < .001 | 162.44 | 0.81 | 0.56 |
| Touches | .69 | .61–.78 | .044 | < .001 | 65.50 | .60 | .78 |
| %Off-Screen | .67 | .59–.76 | .044 | < .001 | .49 | .56 | .72 |

Note. AUC = Area Under the Curve; CI = Confidence Interval; SE = Standard Error. Optimal cutoff, sensitivity, and specificity values are based on the highest Youden’s Index.

**Nonparametric Correlations for All Convergent Validity Analyses**

Supplementary Table 3. Spearman correlation coefficients (and p-values) between VKC-2 scores and Real Kitchen scores (n = 201).

| | Real Kitchen Scores | | | |
|---|---|---|---|---|
| VKC-2 scores | Completion Time | Accomplishment Score | Total Errors | Motor Errors |
| Digital Dexterity | .49* | -.41* | .28* | .12 (p =.118) |
| Time | .56* | -.37* | .44* | .20 (p =.007) |
| Touches | .32* | -.27* | .34* | .28* |
| %Off-Screen | .26* | -.44* | .26* | .15 (p =.057) |

* p < .001 (2-tailed).

Supplementary Table 4. Spearman correlation coefficients (and p-values) between VKC-2 Scores and Cognitive Test Scores (N = 236)

| VKC-2 scores | mKnight-PACC | Executive Function Composite | Episodic Memory Composite | Processing Speed Composite | Language Composite |
|---|---|---|---|---|---|
| Digital Dexterity | -.36* | -.25* | -.31* | -.32* | -.20 (p=.002) |
| Time | -.41* | -.26* | -.36* | -.32* | -.30* |
| Touches | -.17 (p=.010) | -.09 (p=.179) | -.26* | -.07 (p=.314) | -.25* |
| %Off-Screen | -.31* | -.27* | -.33* | -.22* | -.21* |

*p <.001 (2-tailed)

Supplementary Table 5. Spearman correlation coefficients (and p-values) between VKC-2 scores and questionnaires

| | Participant Questionnaires (n = 236) | | | Informant Questionnaires (n = 194) | | |
|---|---|---|---|---|---|---|
| VKC-2 scores | IADL-C | FAQ | ECog | IADL-C | FAQ | ECog |
| Digital Dexterity | .13 (p=.060) | .20 (p=.003) | .15 (p=.029) | .21(p=.003) | .22 (p=.002) | .17 (p=.017) |
| Time | .11 (p=.102) | .24* | .15 (p=.029) | .19 (p=.008) | .23 (p=.001) | .21 (p=.004) |
| Touches | .06 (p=.425) | .22 (p=.001) | .16 (p=.020) | .40* | .14 (p=.047) | .14 (p=.063) |
| %Off-Screen | .24* | .23* | .17 (p=.013) | .21(p=.004) | .25* | .17 (p=.018) |

*p < .001 (2 tailed)

**Test-retest reliability (intraclass correlation coefficients) for participants who reported no cognitive change.**

Intraclass correlation coefficients were run to evaluate test-retest reliability of VKC-2 scores for only participants who reported no cognitive change (n = 123) and are reported below. Cohen's kappa for distractor interactions in this subsample that reported no cognitive change was not statistically significant ($\kappa = 0.14$, $p = .08$), as very few participants in this subsample interacted with distractor objects at session 2 (4.8%; n = 6/123).

Supplementary Table 6. Intraclass correlation coefficients for VKC-2 scores over time for only participants who reported no cognitive change (n = 123)

| Measure | ICC (Average Measures) | 95% CI Lower | 95% CI Upper | F | df1 | df2 | p |
|---|---|---|---|---|---|---|---|
| Digital Dexterity | .831 | .742 | .887 | 6.442 | 122 | 122 | <.001 |
| Time | .836 | .755 | .888 | 6.517 | 122 | 122 | <.001 |
| Touches | .865 | .796 | .910 | 8.027 | 120 | 120 | <.001 |
| %Off-Screen | .709 | .517 | .816 | 3.956 | 120 | 120 | <.001 |
